# Healthy lifestyle for the prevention of post-COVID-19 multisystem sequelae, hospitalization, and death: a prospective cohort study

**DOI:** 10.1101/2024.01.30.24302040

**Authors:** Yunhe Wang, Binbin Su, Marta Alcalde-Herraiz, Nicola L. Barclay, Yaohua Tian, Chunxiao Li, Nicholas J. Wareham, Roger Paredes, Junqing Xie, Daniel Prieto-Alhambra

**Affiliations:** Nuffield Department of Population Health, University of Oxford, Oxford, UK; School of Population Medicine and Public Health, Chinese Academy of Medical Sciences/Peking Union Medical College, Beijing, China; Centre for Statistics in Medicine and NIHR Biomedical Research Centre Oxford, NDORMS, University of Oxford, Oxford, UK; School of Public Health, Tongji Medical College, Huazhong University of Science and Technology, Wuhan, China; Medical Research Council Epidemiology Unit, University of Cambridge, Cambridge, UK; Department of Infectious Diseases & irsiCaixa AIDS Research Institute, Hospital Universitari Germans Trias i Pujol, Catalonia, Spain; Center for Global Health and Diseases, Department of Pathology, Case Western Reserve University School of Medicine, Cleveland, OH, US; Department of Medical Informatics, Erasmus Medical Center University, Rotterdam, Netherlands

## Abstract

**Background:** Post-COVID complications are emerging as a global public health crisis. Effective prevention strategies are needed to inform patients, clinicians and policy makers, and to reduce their cumulative burden. We aimed to investigate whether a habitual healthy lifestyle predated pandemic is associated with lower risks of multisystem sequelae and other adverse outcomes of COVID-19, and whether the potential protective effects are independent of pre-existing comorbidities.

**Methods:** The prospective population-based cohort study enrolled participants with SARS-CoV-2 infection confirmed by a positive polymerase chain reaction test result between March 1, 2020, and March 1, 2022. Participants with no history of the related outcome one year before infection were included and followed up for 210 days. Exposures included ten modifiable healthy lifestyle factors including past or never smoking, moderate alcohol intake (≤4 times week), body mass index <30 kg/m^2^, at least 150 minutes of moderate or 75 minutes of vigorous physical activity per week, less sedentary time (<4 hours per day), healthy sleep duration (7-9 hours per day), adequate intake of fruit and vegetables (≥400 g/day), adequate oily fish intake (≥1 portion/week), moderate intake of red meat (≤4 portions week) and processed meat (≤4 portions week). Outcomes included multisystem COVID-19 sequelae (consisting of 75 diseases/symptoms in 10 organ systems), death, and hospital admission following SARS-CoV-2 infection, confirmed by hospital inpatient and death records. Risk was reported in relative scale (hazard ratio [HR]) and absolute scale (absolute risk reduction [ARR]) during both the acute (the first 30 days) and post-acute (30-210 days) phases of infection using Cox models.

**Findings:** A total of 68,896 participants (mean [SD] age, 66.6 [8.4]; 32,098 women [46.6%]) with COVID-19 were included. A favorable lifestyle (6-10 healthy lifestyle factors; 46.4%) was associated with a 36% lower risk of multisystem sequelae of COVID-19 (HR, 0.64; 95% CI, 0.58-0.69; ARR, 7.08%; 95% CI, 5.98-8.09), compared with unfavorable lifestyle (0-4 factors; 12.3%). Risk reductions were observed across all 10 prespecified organ systems including cardiovascular, coagulation, metabolic and endocrine, gastrointestinal, kidney, mental health, musculoskeletal, neurologic, and respiratory disorders, and general symptoms of fatigue and malaise. This beneficial effect was largely attributable to direct effects of healthy lifestyle, with mediation proportion ranging from 44% to 93% across organ systems. A favorable lifestyle was also associated with lower risk of post-COVID death (HR, 0.59; 95% CI, 0.52-0.66; ARR, 1.99%; 95% CI, 1.61-2.32) and hospitalization (HR, 0.78; 95% CI, 0.73-0.84; ARR, 6.14%; 95% CI, 4.48-7.68). These associations were observed after accounting for potential misclassification of lifestyle factors, and during acute and post-acute infection, in those tested positive in the hospital and community setting, and independent of vaccination status or SARS-CoV-2 variant.

**Interpretation:** Adherence to a healthy lifestyle predated pandemic was associated with substantially lower risk of complications across organ systems, death, and hospitalization following COVID-19, regardless of phases of infection, vaccination status, test setting, and SARS-CoV-2 variants, and independent of comorbidities. These findings illustrate the benefits of adhering to a healthy lifestyle to reduce the long-term adverse health consequences following SARS-CoV-2 infection.

**Research in context:** *Evidence before this study:* We searched PubMed and MEDLINE for articles published between March 1, 2020, and December 1, 2023, using the search terms “healthy lifestyle”, “risk factor”, “post-COVID condition”, “long COVID”, “post-acute sequelae”, “prevention”, “management”, and “treatment”, with no language restrictions. Previous evidence on the prevention and management of long COVID has mainly focused on vaccination and pharmaceutical approaches, including antivirals (e.g., molnupiravir and nirmatrelvir) and other drugs (e.g., metformin). Vaccination before infection or use of antivirals in selected high-risk patients during acute infection only partially mediates the risk of COVID-19 sequelae. Evidence for the non-pharmaceutical prevention strategies are lacking. We identified only two publications on the association between healthy lifestyle and post-COVID condition, and one meta-analysis of the risk factors for long COVID symptoms. A cross-sectional study of 1981 women suggested an inverse association between healthy lifestyle factors and self-reported symptoms following infection of non-Omicron variants, which was mainly driven by BMI and sleep duration. Another study suggested an inverse prospective association between healthy lifestyle prior to infection and post-COVID cardiovascular events. High BMI and smoking are risk factors for long COVID mainly in hospitalized patients. We did not find any study that assessed the association between a composite healthy lifestyle and subsequent post-COVID complications or sequelae across organ systems, hospitalization, and death.

*Added value of this study:* In a prospective, population-based cohort of 68,896 participants with COVID-19, adherence to a healthy lifestyle prior to infection was associated with a substantially lower risk of multisystem sequelae (by 20%-36%), death (by 26%-41%), and hospital admission (by 13%-22%) following COVID-19. The reduced risk of sequelae was evident across 10 prespecified organ systems, including cardiovascular, coagulation and hematologic, metabolic and endocrine, gastrointestinal, kidney, mental health, musculoskeletal, neurologic, and respiratory disorders, as well as general symptoms of fatigue and malaise. The reduced risk of multisystem sequelae, hospitalization, and death associated with a healthy lifestyle was consistently observed across participants, regardless of their vaccination status, disease severity, and major SARS-CoV-2 variants, and largely independent of relevant comorbidities. Adherence to a healthy lifestyle prior to infection was consistently and directly associated with reduced risk of sequelae and other adverse health outcomes following COVID-19.

*Implications of all the available evidence:* The inverse association of healthy lifestyle with multisystem sequelae was even larger than those observed in previous studies of pharmaceutical interventions in non-hospitalized patients. Considering the restricted scope of currently available therapies, such as antivirals (only selected patients at higher risk are qualified during the acute infection) and limited efficacy of vaccination in preventing long COVID, adherence to a healthy lifestyle, in combination with vaccination and, if necessary, potential medications, emerges as practical prevention and care strategies to mitigate the long-term health consequences of SARS-CoV-2 infection. These strategies are of significant clinical and public health importance in reducing the overall burden of post-COVID conditions and improving preparedness for future pandemics.

## Introduction

COVID-19 cases and deaths have decreased globally, yet the long-term health consequences of SARS-CoV-2 infection, termed as *post-COVID-19 conditions* or *long COVID*, are still being managed as a global public health crisis.^1, 2^ These conditions or symptoms can involve pulmonary and multiple extrapulmonary organ systems, and may occur or extend beyond the acute infection of varying severity, with significant impact on daily functioning and quality of life.^3^ Increased risk and burden of cardiovascular, pulmonary, neuropsychiatric, and metabolic disorders were reported during the 6 to 12 months following SARS-CoV-2 infection,^4^ with persistent risk observed for several diseases up to 2 years.^5, 6^

Despite long COVID has been characterized, however, evidence-based strategies for its prevention or treatment are not yet available.^7, 8^ Increasing evidence suggests that vaccination before infection^9, 10^ and use of antivirals^11, 12^ during acute phase only partially mediate the risk of sequelae. Several potential drugs for long COVID are still under investigation without yielding reliable results.^7, 8^ Effective prevention and intervention strategies are needed to inform patients, clinicians and policy makers, and to reduce the cumulative burden of post-COVID conditions.

Modifiable lifestyle factors such as physical activity and healthy diet are potential targets for the prevention of major non-communicable diseases,^13–15^ and are associated with lower risk of severe COVID-19 and related mortality,^16, 17^ possibly through protection against inflammation,^15, 16^ autoimmunity,^18, 19^ and clotting abnormality.^20, 21^ These mechanisms overlap with the hypothesized pathogenesis of long COVID, and other postviral conditions, such as chronic fatigue syndrome.^3^ When examined individually, factors such as smoking and obesity, have been reported to be related to increased risk of post-COVID symptoms mainly in hospitalized patients.^22^ Nevertheless, association between combinations of multiple lifestyle factors, which are known to interact synergistically,^23^ and risk of COVID-19 sequelae across multiple organ systems remains unclear.

This major knowledge gap should be urgently addressed to inform the prevention and care strategies of long COVID. Based on a large-scale, prospective population-based cohort, we evaluated the relationship between composite healthy lifestyle (including 10 modifiable factors) predated pandemic and subsequent risk of COVID-19 sequelae in 10 organ systems, death and hospital admission, while considering the phase of infection (acute or post-acute), severity of infection (tested positive in community/outpatient setting vs inpatient setting), vaccination status (fully vaccinated vs unvaccinated or partially vaccinated), and variants (alpha [B.1.1.7], delta [B.1.617.2], vs omicron [B.1.1.529]), that differ in transmissibility, disease course, and disease severity.^24^

## Methods

### Data Sources and Study Cohorts

UK Biobank is a large-scale population-based prospective cohort study with deep phenotyping and genomic data, as detailed elsewhere.^25^ Briefly, between 2006 to 2010, over 500,000 individuals aged 40-69 years were recruited from 22 assessment centers across the United Kingdom at baseline, with collection of socio-demographic, lifestyle and health-related factors, a range of physical measures, and blood samples.^25^ Follow-up information is obtained by linking health and medical records, including national primary and secondary care, disease and mortality registries,^26^ with validated reliability, accuracy and completeness.^27^ To identify cases of SARS-CoV-2 infection, polymerase chain reaction (PCR)-based test results were obtained by linking all participants to the Public Health England’s Second Generation Surveillance System, with dates of specimen collection and healthcare settings of testing.^28^ Outbreak dynamics were validated to be broadly similar between UK Biobank participants and the general population of England.^28^

In this study, we enrolled participants who were alive by March 1, 2020 and had a positive SARS-CoV-2 PCR test result between March 1, 2020 (date of the first recorded case in the UK Biobank), and March 1, 2022, with the date of first infection considered as index date (*T_0_*). The major prevalent variants during the study period included wildtype, Alpha (B.1.1.7), Delta (B.1.617.2), and Omicron (B.1.1.529 BA.1). The calendar periods of dominant variants in the UK were based on pandemic data from the Office for National Statistics.^24^ Participants with missing data on study exposures and covariates of interest at baseline were further excluded. All participants included in this study provided written informed consent at recruitment. This study followed the Strengthening the Reporting of Observational Studies in Epidemiology (STROBE) reporting guidelines and received ethical approval from the UKBB ethics advisory committee. Study design, cohort construction, and timeline are provided in **Supplementary Fig. 1**.

### Lifestyle factors

Ten prespecified potentially modifiable lifestyle factors were assessed, including smoking, alcohol consumption, body mass index (BMI), physical activity, sedentary time, sleep duration, intake of fruit and vegetable, intake of oily fish, intake of red meat, and intake of processed meat. Selection and categorization of lifestyle factors was based on literature review, previous knowledge, and UK national health service guidelines.^29, 30^ Multiple lifestyle factors were measured by validated questionnaire for all participants at baseline recruitment. Part of participants took part in up to two further touchscreen interviews with lifestyle and health-related factors similarly measured. There were generally stable responses to lifestyle factors between baseline assessment and the latest repeat assessment (median time difference from baseline, 8 years) as shown in **Supplementary Fig. 2**. Detailed definitions on measurement and classification of lifestyle factors are provided in **Supplementary Table 1**. Briefly, healthy lifestyle components including past or never smoker, moderate alcohol intake (≤4 times week), BMI <30 kg/m^2^, at least 150 minutes of moderate or 75 minutes of vigorous physical activity per week, less sedentary time (<4 hours per day), healthy sleep duration (7-9 hours per day), adequate intake of fruit and vegetables (≥400 g/day), adequate oily fish intake (≥1 portion/week), moderate intake of red meat (≤4 portion week) and processed meat (≤4 portion week) were defined, in accordance with previous evidence or UK national health service guidelines.^29, 30^

A binary variable was created for each of the 10 factors, with 1 point assigned for those meeting the healthy criteria and 0 otherwise. A composite lifestyle score was then calculated for each participant by summing the total number of healthy lifestyle factors, ranging from 0 to 10. Based on the composite score, participants were classified into three lifestyle categories: unfavorable (0-5), intermediate (6-7), and favorable (8-10). The lifestyle score was also used as a continuous variable of number of healthy lifestyle factors. Similar methods of defining lifestyle score have been used in the same UK Biobank cohort^31^ as well as external cohorts.^14, 32^ Distributions of lifestyle score and categories are provided in **Supplementary Table 2.**

### Outcomes

The outcomes after COVID-19 were prespecified, including a set of multisystem sequelae, death, and hospital admission following the SARS-CoV-2 infection. The multisystem sequelae were selected and defined based on previous evidence of the long COVID, including 75 systemic diseases or symptoms in 10 organ systems: cardiovascular,^33^ coagulation and hematologic,^33^ metabolic and endocrine,^34^ gastrointestinal,^35^ kidney,^36^ mental health,^37^ musculoskeletal,^38^ neurologic,^38^ and respiratory disorders,^9, 11, 12^ and general symptoms of fatigue and malaise.^3, 4, 39, 40^ Detailed definitions of multisystem sequelae are listed in the **Supplementary Table 3**. Outcomes were identified as follows: individual sequela from the hospital inpatient ICD-10 (*International Classification of Diseases 10th Revision*) diagnosis codes, deaths from the records of national death registry, and hospital admission from hospital inpatient data from the Hospital Episode Statistics. Incident outcomes were assessed in participants with no history of the related outcome within one year before the date of the first infection.

As SARS-CoV-2 infection has been associated with both multisystem manifestations during its acute phase and with sequelae during its post-acute phase,^6, 40^ we conducted analyses stratified by phase of infection. We reported risk of each outcome during the acute phase (*T_0_* to *T_0_* + 30d), post-acute phase (*T_0_* + 30d to *T_0_* + 210d), and overall period following infection (*T_0_* to *T_0_* + 210d) to reflect the full spectrum of post-COVID conditions. The end of follow-up for the overall cohort was September 30, 2022, with the maximum follow-up period censored to 210 days.

### Covariates

We prespecified a list of covariates for adjustment or stratification based on literature review and prior knowledge: sociodemographic characteristics including age, sex, education level (mapped to the international standard for classification of education), index of multiple deprivation (IMD, a summary measure of crime, education, employment, health, housing, income, and living environment),^41^ and race and ethnicity; and infection related factors including healthcare settings of the testing (community/outpatient vs inpatient setting as proxy of severity of infection), COVID-19 vaccination status, and SARS-CoV-2 variants.

### Statistical analysis

Baseline characteristics of the overall cohort of participants with SARS-CoV-2 infection and by composite healthy lifestyle categories were reported as mean and standard deviation or frequency and percentage, when appropriate. Multivariable cox proportional hazard (PH) model was used to assess the association between composite healthy lifestyle and risk of multisystem sequelae (composite or by organ systems), death, and hospital admission, with adjustment for age, sex, ethnicity, education level, and IMD. PH assumption across lifestyle categories was tested by Schoenfeld residuals with no violations observed for outcomes. Hazard ratio (HR) and absolute risk reduction (ARR, difference in incidence rate between lifestyle groups per 100 persons during the corresponding follow-up period) were estimated from the Cox model. We also assessed the association between individual lifestyle factor instead of composite categories (each component as a categorical variable with or without mutual adjustment for others, or the number of factors as continuous variables) and risk of outcomes.

We conducted causal mediation analysis^42, 43^ to quantify the extent to which the habitual healthy lifestyle may affect COVID-19 sequelae through the potential pathway of relevant pre-infection medical conditions (mediator), with proportion of direct and indirect effects estimated by quasi-Bayesian Monte Carlo methods with 1000 simulations for each. Detailed modelling procedures and a directed acyclic graph are provided in eMethods in Supplement.

We examined the association between composite healthy lifestyle and overall risk of multisystem sequelae in prespecified clinical subgroups by demographic and infection-related factors. The demographic factors included age (≤65 and >65 years), sex (male and female), and ethnicity (White and other ethnic groups). As the risk profile of COVID sequelae was related to vaccination and severity of infection, and may change with the evolving pandemic, infection-related factors including vaccine status (no or one-dose partial vaccination and two-dose full vaccination), test setting (inpatient and outpatient or community), dominant variants during the study period (wildtype, Alpha, Delta, and Omicron BA.1) were assessed. Multiplicative interactions between the composite healthy lifestyle and the stratification variables were tested, with *P*-value reported.

We conducted multiple sensitivity analyses to assess the robustness of primary findings. First, to reflect the multisystem and potentially comorbid nature of COVID sequelae, accounting for both the number of sequelae by an individual and the relative health impact of each sequela. Weights based on Global Burden of Disease study data and methodologies for general diseases and long COVID were assigned to each sequela (**Supplementary Table 1**).^44, 45^ The weighted score was calculated for each participant by summing the weights of all incident sequelae during the follow-up period. Zero inflated Poisson regression was then used to calculate relative risk (RR), with follow-up time set as the offset of the model and adjustment for covariates. Second, to further account for potential reverse causality and more accurately define incident cases, extending the washout period for outcomes from one year to two years. Third, defining events of post-acute sequelae 90 days after infection (follow-up period *T_0_* + 90d to *T_0_* + 210d), instead of 30 days in the main analyses. The adjustment was made as there is no uniform definition for long COVID, which is currently described as conditions occurring 30-90 days after infection in existing guidelines.^46^ Fourth, restricting the identification of outcomes to the first three ICD diagnoses, which are the main causes for each hospital admission.

Fifth, we conducted quantitative sensitivity analysis to adjust for changes in lifestyle factors over time since the baseline assessment. We used odds ratios to quantify associations and assumed a sensitivity and specificity of 90% for each lifestyle component (eMethods in Supplement).

Statistical significance was determined by a 95% confidence interval (CI) that excluded 1 for ratios and 0 for rate differences. All analyses and data visualizations were conducted using R statistical software (version 4.2.2).

## Results

### Baseline Characteristics

Out of 472,977 eligible UK Biobank participants, 68,896 participants with a positive SARS-CoV-2 test result between March 1, 2020 and March 1, 2022 were included in the current study. The demographic and health characteristics of the eligible participants, and participants with COVID-19 overall and by healthy lifestyle category are provided in **Table 1** Of the COVID-19 cohort, the mean (SD) age was 66.6 (8.4) years, 53.4% were male and 82.1% were White. For composite healthy lifestyle prior to the infection, 12.3% followed an unfavorable lifestyle, 41.3% followed an intermediate lifestyle, and 46.4% followed a favorable lifestyle. The median [IQR] number of healthy lifestyle factors participants engaged in was 7 [6-8]. For prespecified COVID-19 sequalae, 5.5% and 7.8% had sequelae in at least one organ system during the acute and post-acute phase of infection, respectively.

**Table 1.**
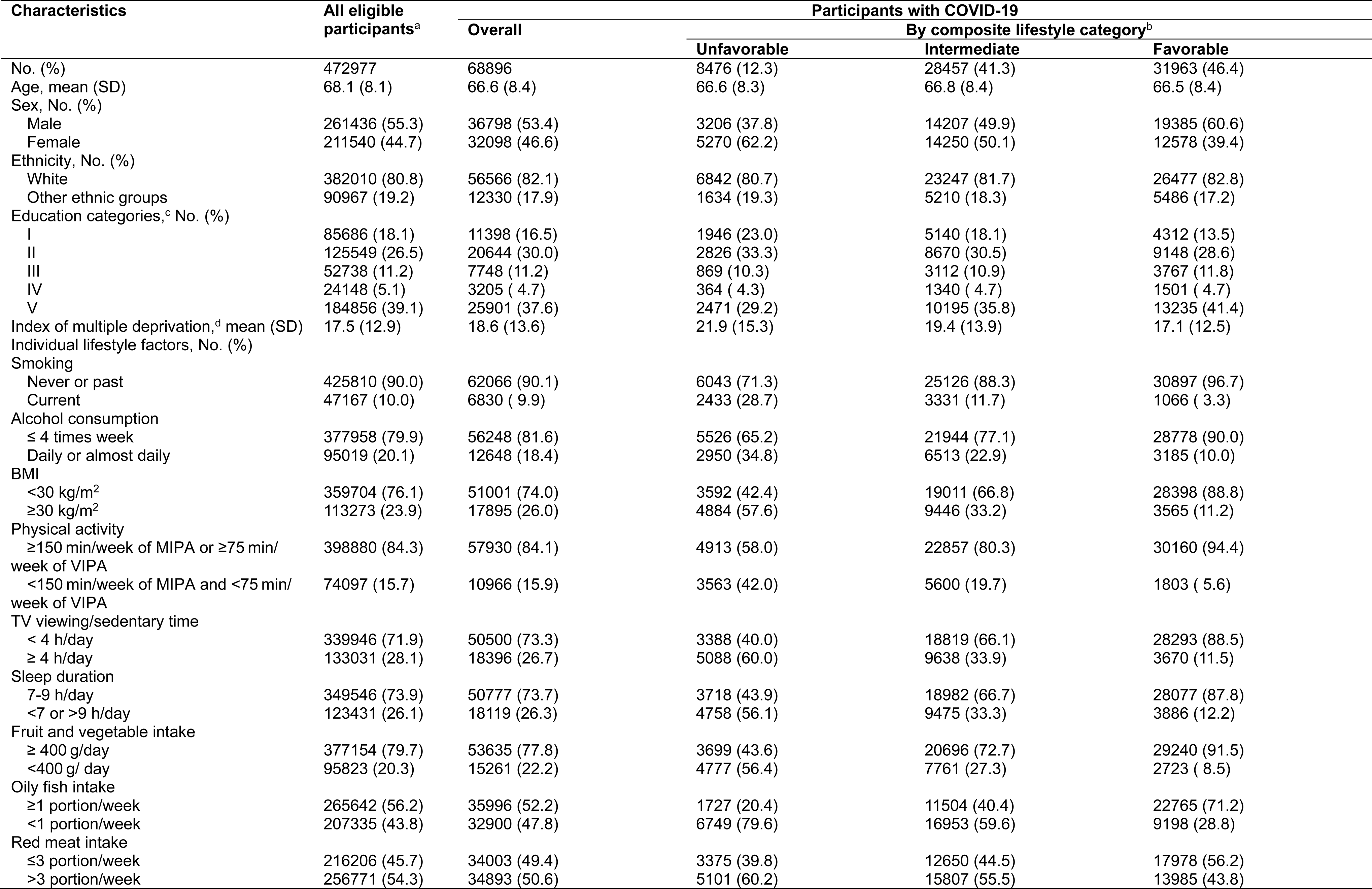
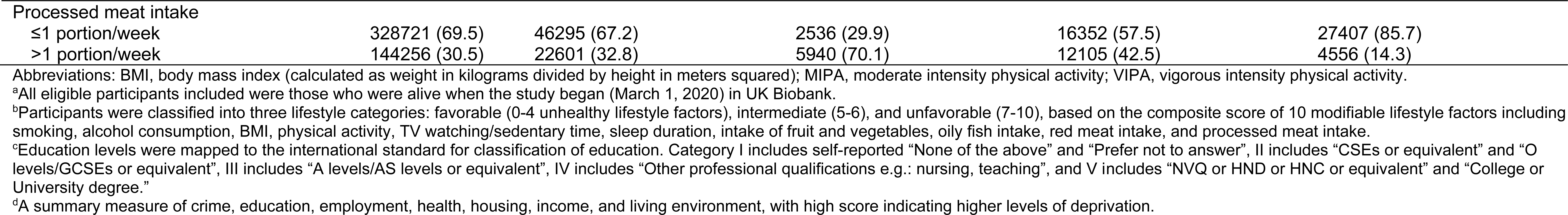
Baseline characteristics of eligible participants, and COVID-19 cohort overall and by lifestyle category.

### Risk of multisystem sequelae

Overall, the risk of multisystem COVID-19 sequelae decreased monotonically across healthy lifestyle categories during both the acute and post-acute phases of infection. Compared with those with an unfavorable lifestyle, participants with an intermediate (HR, 0.80; 95% CI, 0.74-0.87; ARR at 210 days, 3.89%; 95% CI, 2.56-5.12) and favorable (HR, 0.64; 95% CI, 0.58-0.69; ARR at 210 days, 7.08%; 95% CI, 5.98-8.09) lifestyle were at significantly lower risk of multisystem sequelae of COVID-19 (**Figure 1**A and eFigure 3 in **<u>Supplement</u>**), with similar trends observed in both the acute and post-acute phases of COVID-19 (**Figure 1**B). The number of healthy lifestyle factors (range, 0-10) was associated with risk of sequelae in a dose-dependent manner (**Figure 1**A).

**Figure 1.**
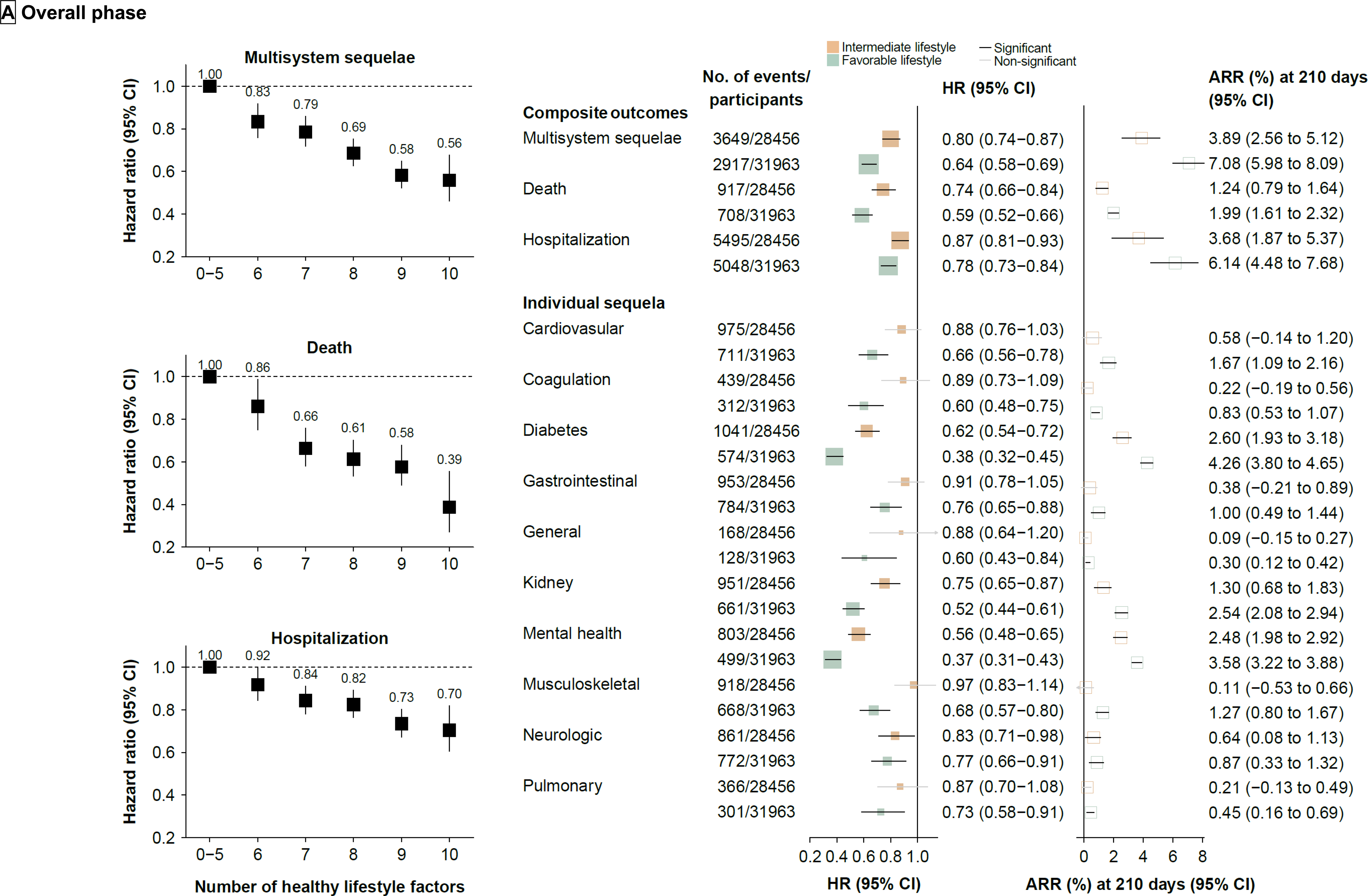
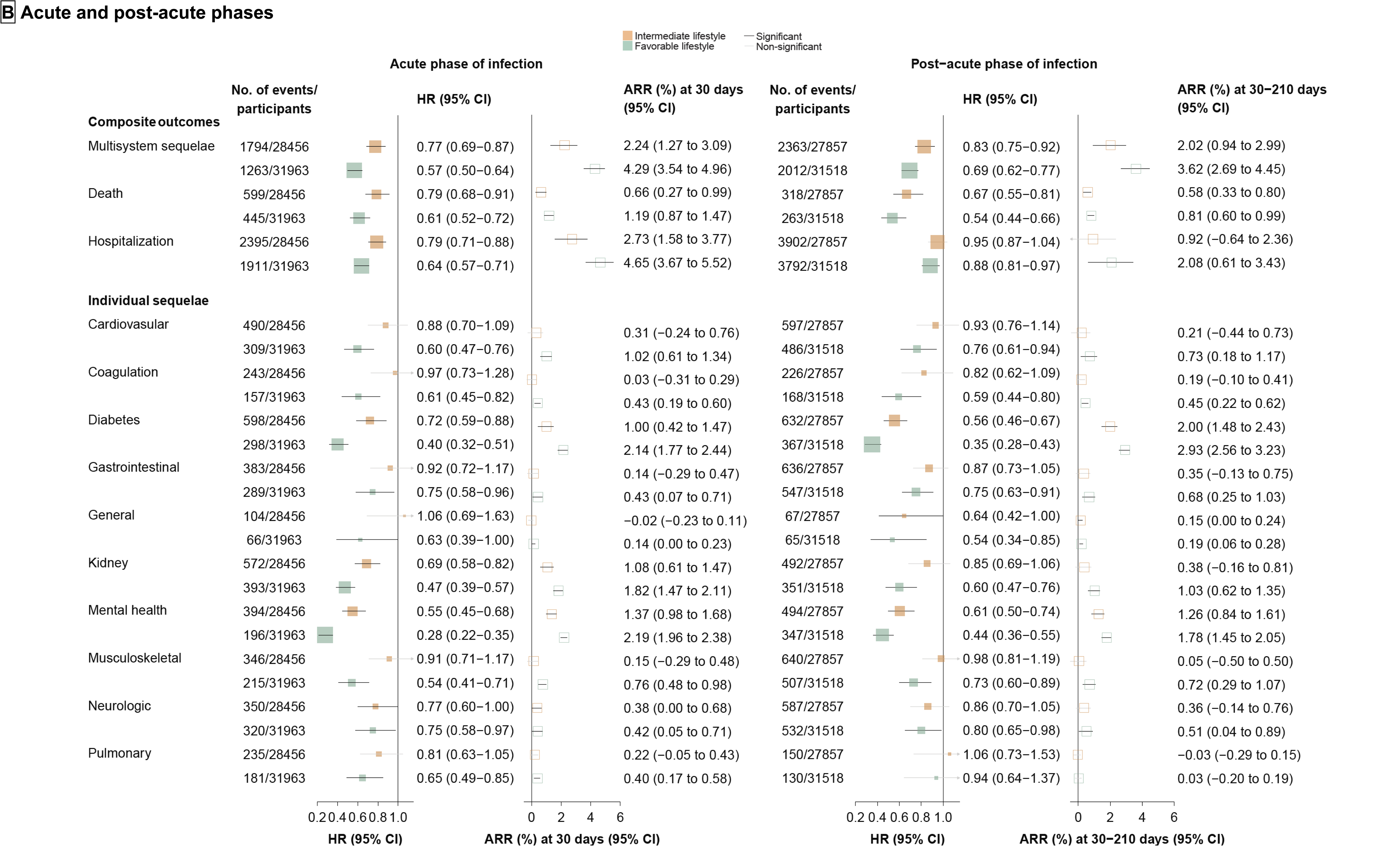
Association of healthy lifestyle with multisystem sequelae of COVID-19, death, and hospital admissions, during overall, acute, and post-acute phases of SARS-CoV-2 infection. A, Healthy lifestyle (composite or number) and risk of multisystem sequelae (composite or by organ systems), death, and hospitalization during the overall phase (0-210 days) of SARS-CoV-2 infection. B, Composite healthy lifestyle and risk of multisystem sequelae, death, and hospitalization during the acute phase (first 30 days) and post-acute (30-210 days) phases of SARS-CoV-2 infection. Adjusted RRs and 95% CI are presented for composite multisystem sequelae, and adjusted HRs and 95% CIs for other outcomes. Absolute risk reduction per 100 persons at 210 days, 30 days, and 30-210 days and 95% CI were calculated. Solid square represents HRs with the area inversely proportional to the variance of the log HR. Hollow square represents ARR. The horizontal lines indicate 95% CIs, with black line representing statistically significant results and grey line representing non-significant results. Intermediate lifestyle category are in orange, favorable lifestyle category in green.

The inverse associations with multisystem sequelae were largely attributable to the direct protective effect of healthy lifestyle (proportion of direct effect on any sequela: 71%), with proportion of direct effect ranging from 44% to 93% across organ systems (**Figure 2**A). Pre-infection medical conditions were associated with substantially increased risk of COVID-19 sequelae, particularly history of cardiovascular diseases, diabetes and mental disorders (**Figure 2**B).

**Figure 2.**
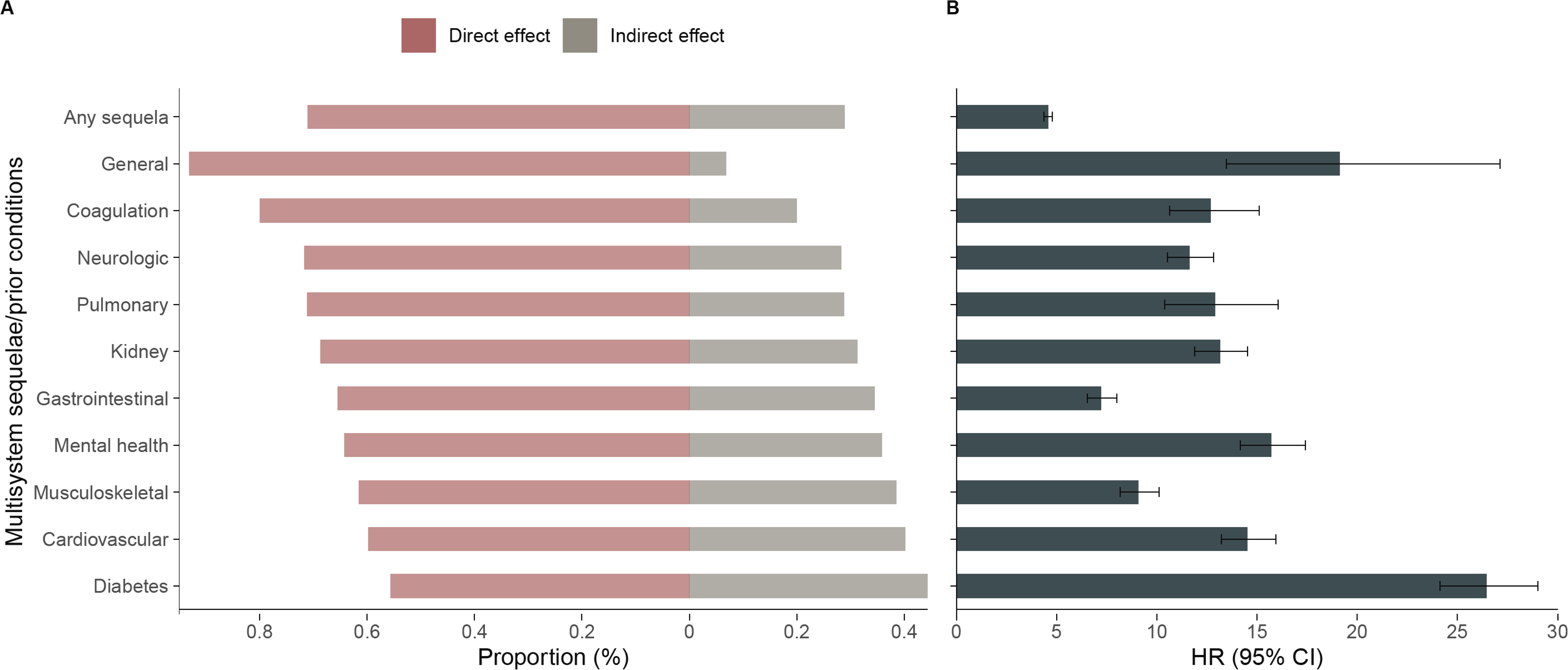
Direct and indirect effects of healthy lifestyle, and association of pre-infection medical conditions with multisystem sequelae of COVID-19. A, Proportion of the direct and indirect effect of healthy lifestyle on multisystem sequelae (Intermediate/favorable vs unfavorable lifestyle). Direct association were accounted for pre-infection medical conditions (mediator), identified as any relevant event recorded between baseline measurement and infection date. B, Association of corresponding pre-infection medical conditions with risk of sequelae following SARS-CoV-2 infection. Outcomes were ascertained 0-210 days after SARS-CoV-2 infection.

**Figure 3.**
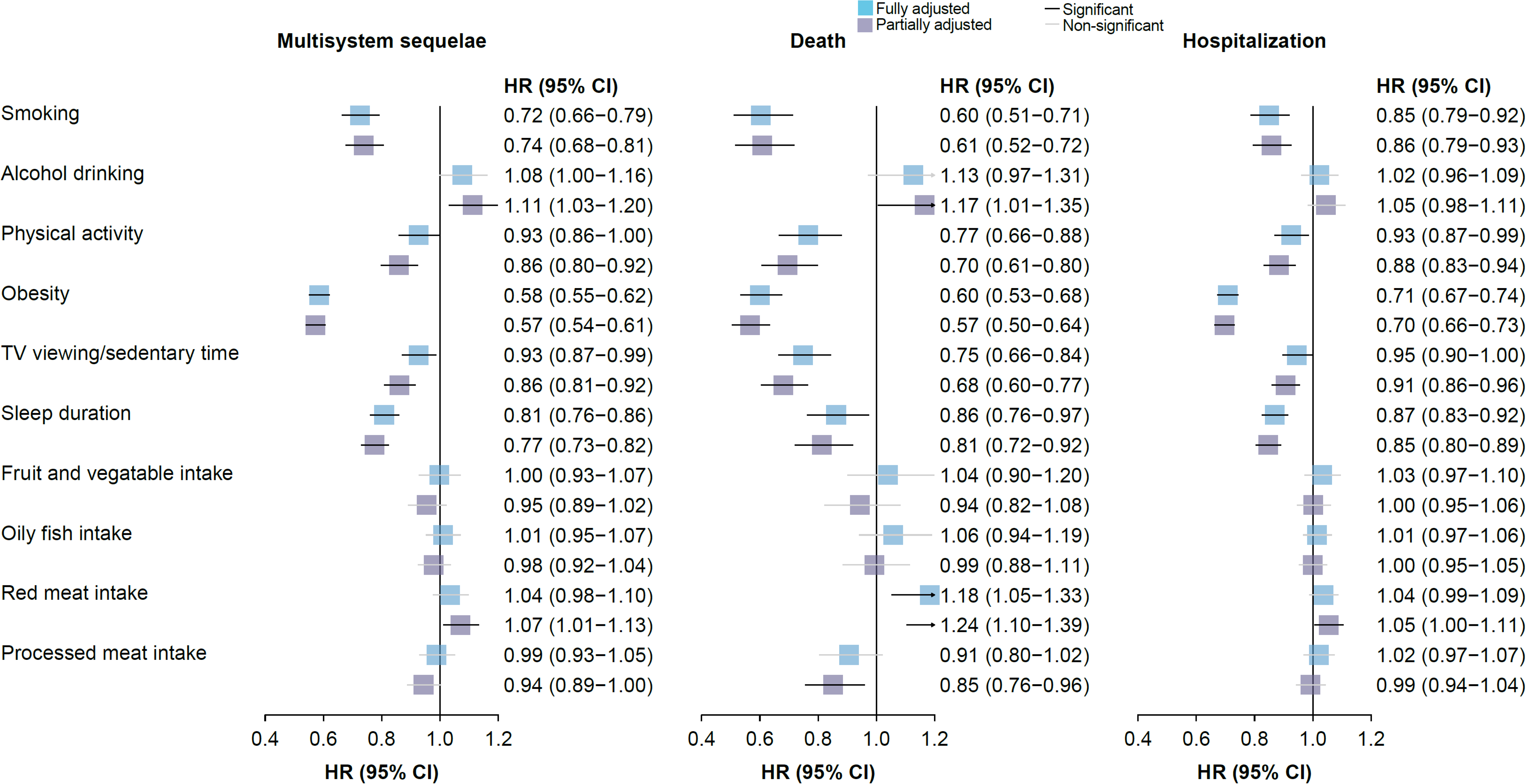
Association of individual components of composite healthy lifestyle with multisystem sequelae, death, and hospitalization. Blue square represents risk estimates from models fully adjusted for age, sex, education level, ethnicity, IMD, and mutually for all lifestyle factors. Purple square represents risk estimates from models partially adjusted for age, sex, education level, ethnicity, and IMD. The horizontal lines indicate 95% CIs, with black line representing statistically significant results and grey line representing non-significant results.

For individual components of the healthy lifestyle, each of the 10 studied behavioral and dietary factors was associated with a lower or non-differential risk of sequelae, with smoking, physical activity, obesity and sleep duration contributing most (**Figure 3**).

### Risk of death and hospitalization

Adherence to a healthy lifestyle was associated with lower risk of death and hospitalization following COVID-19 during both the acute and post-acute phases of infection. Compared with those with an unfavorable lifestyle, participants with a favorable lifestyle were at significantly lower risk of death (HR, 0.59; 95% CI, 0.52-0.66; ARR at 210 days, 1.99%; 95% CI, 1.61-2.32) and hospitalization (HR, 0.78; 95% CI, 0.73-0.84; ARR at 210 days, 6.14%; 95% CI, 4.48-7.68) following COVID-19 (**Figures 1**A and eFigure 3 in **<u>Supplement</u>**), with similar trend observed in both the acute and post-acute phases (**Figures 1**B). Each of 10 lifestyle factors was associated with a lower or non-differential risk of death and hospitalization (**Figure 3**).

### Risk of system-specific sequelae

Compared with those following an unfavorable lifestyle, participants with a favorable lifestyle had a significantly lower risk of sequelae in all 10 organ systems examined, including cardiovascular, coagulation and hematologic, metabolic and endocrine, gastrointestinal, kidney, mental health, musculoskeletal, neurologic, and respiratory disorders, as well as general symptoms of fatigue and malaise, with overall HRs ranging from 0.38 to 0.76 (**Figure 1**A and **Figure 1**B). The associations with intermediate lifestyle were consistently in the same protective direction across system-specific sequelae (**Figure 1**A and **Figure 1**B). Similar trend were observed in both the acute and post-acute phases (**Figure 1B**).

### Risk of outcomes by subgroups

The inverse associations between healthy lifestyle and risk of multisystem sequelae, death, and hospitalization held across the different subgroups of clinical interest, including those by age, sex, and ethnicity, vaccine status, test setting, and variants of infection (**Table 2**). The reduced risk of outcomes was observed in participants who received two doses of vaccine (breakthrough infection) and those who were unvaccinated or partially 1-dose vaccinated (non-breakthrough infection). The reduced risk was evident in participants tested positive in inpatient setting and in those tested positive in community/outpatient settings. The reduced risk was consistently observed across predominant variants of SARS-CoV-2 infection during study period, including wild-type, Alpha, Delta, and Omicron BA.1. Notably, composite healthy lifestyle was associated with decreased risk of outcomes following infection of Omicron variant, which remains currently dominant variant worldwide. No significant interaction was observed in any of the subgroups across outcomes, except for age, ethnicity, and vaccination. The observed association between favorable lifestyle and a reduced risk of outcomes was more evident for people aged <65, in white subjects, and in those fully vaccinated (for mortality only).

**Table 2.**
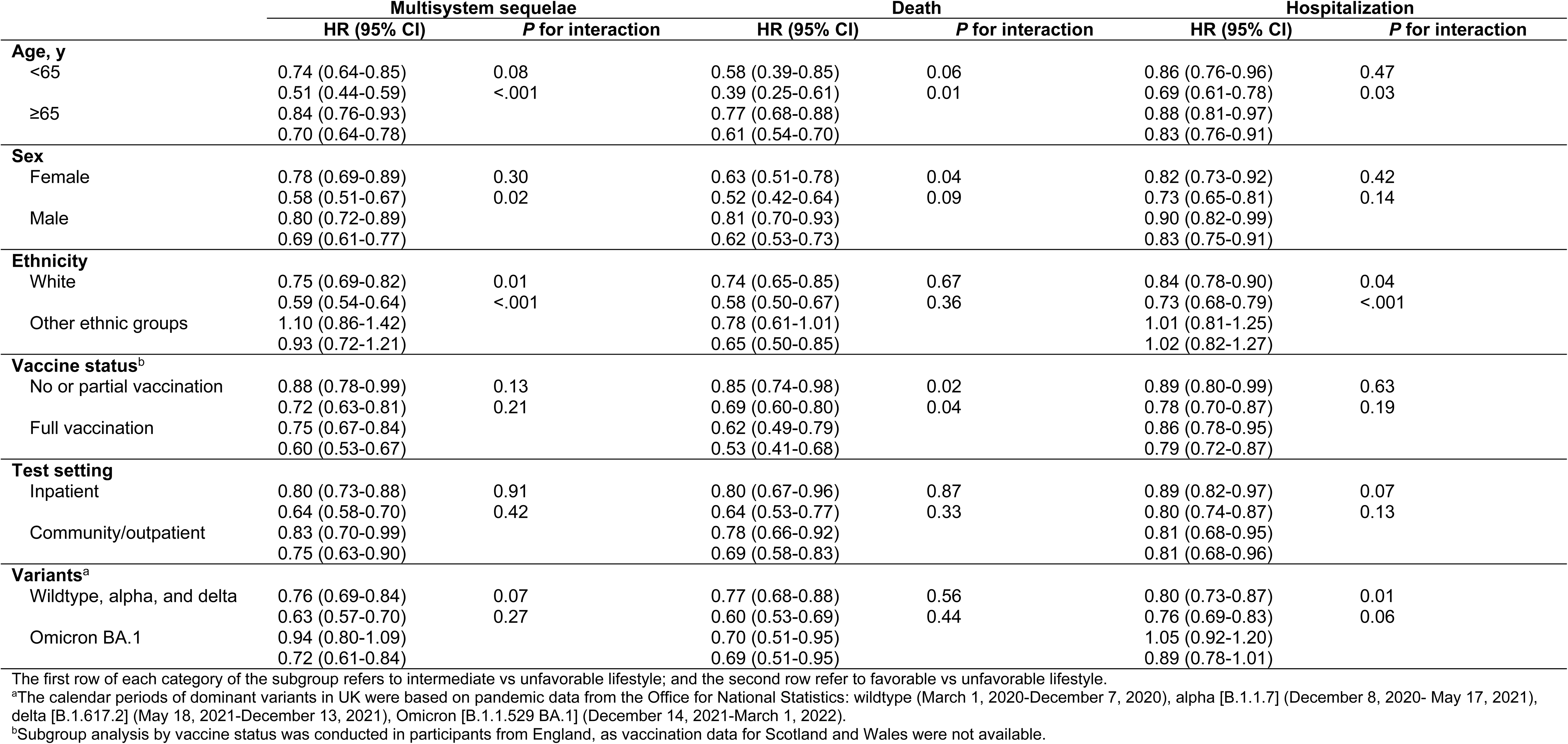
Association of composite healthy lifestyle with multisystem sequelae, death, and hospitalization following SARS-CoV-2 infection in key clinical subgroups.

### Sensitivity analyses

A similar pattern of associations was observed in multiple sensitivity analyses, including assigning weight to individual sequela and using zero inflated Poisson regression to estimate the association, excluding participants with history of related outcomes in the past two years rather than one year, defining post-acute outcomes 90 days after infection rather than 30 days, and restricting the identification of outcomes to the first three ICD diagnoses (eTable 4 in **<u>Supplement</u>**). Stronger associations were identified after accounting for potential misclassification of lifestyle factors, suggesting that the observed associations may have been underestimated (eTable 4 in **<u>Supplement</u>**).

## Discussion

Based on a large, prospective population-based cohort, this study provides a comprehensive assessment of the health effects of multiple lifestyle factors on a systematic range of disease outcomes following COVID-19. Adherence to a healthy lifestyle prior to infection was associated with significantly lower risk of COVID-19 multisystem sequelae, death, and hospital admission, during both the acute and post-acute phases of SARS-CoV-2 infection. The reduced risk was evident across 10 prespecified organ systems, including cardiovascular, coagulation and hematologic, metabolic and endocrine, gastrointestinal, kidney, mental health, musculoskeletal, neurologic, and respiratory disorders, as well as general symptoms of fatigue and malaise. The reduced risk of multisystem sequelae associated with a healthy lifestyle was consistently observed across participants, regardless of their vaccination status (unvaccinated/partially vaccinated or fully vaccinated), disease severity (testing positive in community/outpatient settings or inpatient settings), and major SARS-CoV-2 variants, including Omicron variants, whose subvariants are currently dominant. Moreover, the benefits of healthy lifestyle on sequelae were largely independent of pathways related to pre-existing disease conditions. Overall, the findings suggest that adherence to a healthy lifestyle prior to infection was consistently and directly associated with reduced risk of adverse health outcomes following COVID-19.

We found that a favorable lifestyle, in comparison with an unhealthy one, was associated with a 36% lower risk of multisystem sequelae, a 41% lower risk of death, and a 22% lower risk of hospitalization, which corresponded to an absolute risk reduction of 7.08, 1.99, and 6.14 fewer cases per 100 people at 210 days after infection. This association was even larger than those observed in previous studies of pharmaceutical interventions in non-hospitalized patients, which reported a 14% risk reduction in post-acute sequelae at 180 days for vaccination before infection, and 14% and 26% risk reductions at 180 days for the use of molnupiravir and nirmatrelvir during acute phase of infection, respectively.^9, 11, 12^ It is important to note that participants with breakthrough infection were still at risk of sequelae compared with those without infection.^9^ In addition, only selected patients at risk of progression to severe COVID-19 are qualified for antivirals during the acute infection,^11, 12^ and their benefit-risk profile in wider population with milder infection, or when used during the post-acute stage, remains unclear. These previous findings highlighted the restricted scope of currently available therapies and limited efficacy of vaccination in preventing long COVID.^9, 11, 12, 46^ Our results are consistent with a cross-sectional study of 1981 women suggesting an inverse association between composite healthy lifestyle (mainly driven by BMI and sleep duration) and self-reported symptoms following infection of non-Omicron variants.^32^ However, outcomes purely based on self-report symptoms are less clinically relevant and the inclusion of only women may limit the generalization of findings to other populations and settings.

The mechanisms underlying the benefit of adhering to healthy lifestyle for the alleviation of sequelae are likely multifaceted. Previous research has established causal links between several individual lifestyle factors, such as smoking, obesity, and physical inactivity, and increased susceptibility and severity in relation to COVID-19.^47^ Smoking and high BMI were also risk factors for long COVID symptoms mainly in hospitalized patients.^22^ Indeed, we observed that these factors and additionally sleep duration and sedentary behavior were significant contributors to the reduced risk of sequalae. However, these pathways are unlikely to fully explain the protective effects conferred by healthy lifestyle. In our study, all participants had a confirmed infection, and the protective associations persisted even among those who were hospitalized. In addition, it has been suggested that individuals with an unhealthy lifestyle are more likely to have prevalent chronic conditions, such as cardiovascular diseases and diabetes—which are strong risk factors for severe COVID-19—and are therefore more volunerable to post-acute complications. Through mediation analysis, our study supported this hypothesis and, for the first time, further demonstrated that the healthy lifestyle’s direct protection accounts for the majority of the overall associations with COVID-19 sequelae.

Although our findings align with previous evidence on the broader benefits of healthy lifestyle on chronic disease prevention and life expectancy,^13–15^ this potentially beneficial effect should not be interpreted as changing behaviors around the time of acute infection or during post-acute infection. The current practical guide recommends that patients without long COVID should gradually and safely return to pre-infection physical activity when appropriate, although direct evidence is lacking.^48^ Previous study also characterized long COVID as a multifactorial condition determined by pathogen, host response, external pandemic-associated factors, and supported a multidisciplinary treatment including both pharmacological and rehabilitation approaches, but also social and welfare support to promote healthy lifestyle habits.^49, 50^ Future research is warranted to assess the effect of composite lifestyle interventions in prevention of long COVID or alleviating associated symptoms among patients with long COVID. Adherence to a healthy lifestyle, in combination with vaccination and, if necessary, potential medications, may be a viable and practical approach to further reduce the long-term health consequences of SARS-CoV-2 infection. These strategies hold significant public health and scientific importance in mitigating the overall burden of post-COVID complications and enhancing preparedness for future pandemics.

### Limitation

This study has several limitations. First, the UK Biobank participants are likely to be older and healthier than the general population of the UK and are mostly of European ancestry, which may limit the generalizability of study findings. Despite the relative risk were largely not influenced, the absolute risk should be interpreted with caution. Second, residual confounding and reverse causality cannot be ruled out in this observational study. Third, we assumed the baseline lifestyle unchanged over years until the time of infection, which may be subject to exposure misclassification and underestimated any genuine associations. However, reassuringly, there was high consistency of lifestyle measures between cohort baseline and repetitive interviews after a median of 8 years, and consistent associations were observed after accounting for potential changes in lifestyle factors. Forth, as sequelae outcomes were based on inpatient records, milder long COVID symptoms were less likely to be detected. Fifth, given the potential non-linear effects of lifestyle factors, such as alcohol consumption, caution is warranted when interpreting associations between binarized lifestyle factors and outcomes. Finally, despite we included a range of sequelae across organ systems, it is difficult to link these outcomes directly to the infection, especially given the lack of consensus standard for diagnosis of long COVID. This limitation is applied to all related studies in the field.^33–40, 51^ Nevertheless, the sequelae prespecified were most relevant to long COVID based on prior evidence, with increased risk and burden consistently reported beyond the acute phase in the currently largest electronic dataset,^33–39^ the same UK Biobank,^51^ and other nationwide cohorts.^40, 51^

## Conclusion

Adherence to a healthy lifestyle predated pandemic was associated with substantially lower risk of sequelae across organ systems, death, and hospitalization following COVID-19, regardless of phases of infection, vaccination status, test setting, and SARS-CoV-2 variants, and independent of relevant comorbidities. These findings suggest the benefit of population adhering to a healthy lifestyle to reduce the potential long-term adverse health consequences of COVID-19.

## Data Availability

All data produced in the present study are available upon reasonable request to the authors

https://www.ukbiobank.ac.uk/

## Acknowledgements

This study was based on data from UK Biobank. All participants provided written informed consent at the UK Biobank cohort recruitment. This study received ethical approval from UK Biobank Ethics Advisory Committee (EAC) and was performed under the application of 65397. Mr Wang is funded through the Clarendon Fund Scholarship. Dr Xie is funded through Jardine-Oxford Graduate Scholarship and a titular Clarendon Fund Scholarship. The research was partially supported by the Oxford National Institute for Health and Care Research (NIHR) Biomedical Research Centre. Dr Prieto-Alhambra is funded through an NIHR Senior Research Fellowship (grant SRF-2018-11-ST2-004). The funding organizations had no role in the design and conduct of the study; collection, management, analysis, and interpretation of the data; preparation, review, or approval of the manuscript; and decision to submit the manuscript for publication. The views expressed in this publication are those of the author(s) and not necessarily those of the NHS, the NIHR or the Department of Health.

## Author contributions

Drs Wang and Xie had full access to all the data in the study and take responsibility for the integrity of the data and the accuracy of the data analyses. *Concept and design*: Wang and Xie. *Acquisition, analysis, or interpretation of data*: Wang, Xie, and Prieto-Alhambra. *Drafting of the manuscript*: Wang. *Critical revision of the manuscript for important intellectual content*: All authors. *Statistical analysis*: Xie. *Obtained funding*: Prieto-Alhambra. *Administrative, technical, or material support*: Prieto-Alhambra. *Supervision*: Nicholas J. Wareham and Prieto-Alhambra.

## Supplemental Online Content

### eMethods

#### Causal mediation analysis

We used causal mediation analysis to specifically evaluate the extent to which a habitual healthy lifestyle may affect an COVID-19 sequelae outcome through a potential pathway of relevant medical conditions (mediator) prior to the infection. The mediator of interest was defined as relevant events that occurred between the baseline lifestyle assessment and the date of COVID-19 diagnosis. The statistical analysis was executed in a four-step process: (1) we first fitted a logistic regression model with the mediator as dependent variables and all covariates from the primary analysis as independent variables. (2) another logistic regression model was then fitted with multisystem sequelae as the dependent variable and the corresponding mediator along with other covariates as independent variables. (3) the direct effects (DE), indirect effects and their 95% confidence intervals (IE) were estimated using quasi-Bayesian Monte Carlo methods with 1,000 simulations for each. (4) finally, the mediation proportion was computed as IE/(IE+DE)*100.

Directed acyclic graph (DAG) for identifying confounding variables:

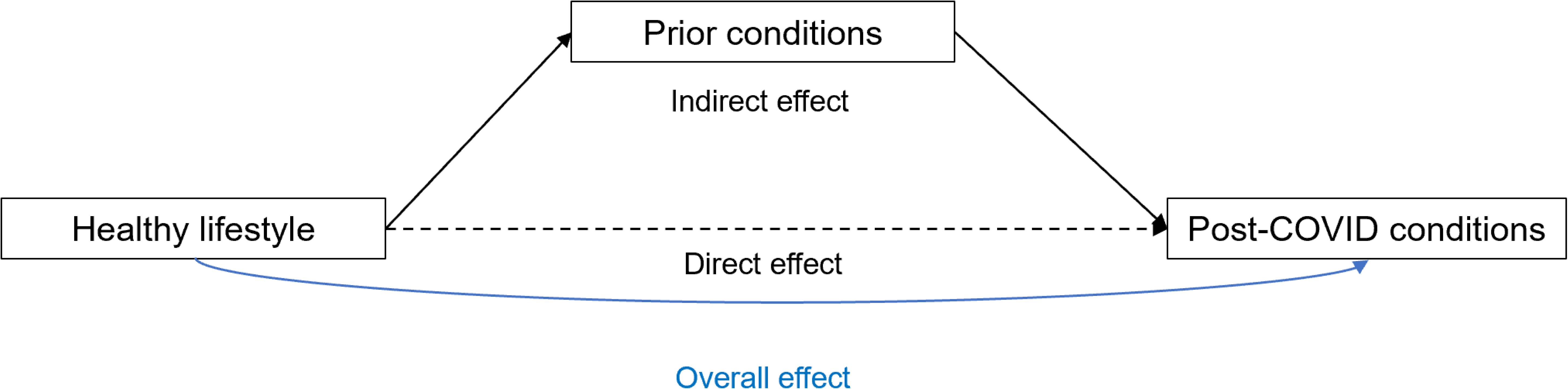

#### Quantitative sensitivity analysis

Quantitative sensitivity analysis was used to adjust for changes in lifestyle factors over time since the baseline assessment. We used a standard algebraic approach that recalculates the expected cell frequencies for a correctly classified 2 x 2 contingency table based on the observed frequencies in a misclassified table. We then derived the bias-adjusted odds ratio using the formula A*D/B*C, assuming non-differential sensitivity and specificity both at 0.9. The equations used to calculate the expected true data, considering exposure misclassification, are as follows:

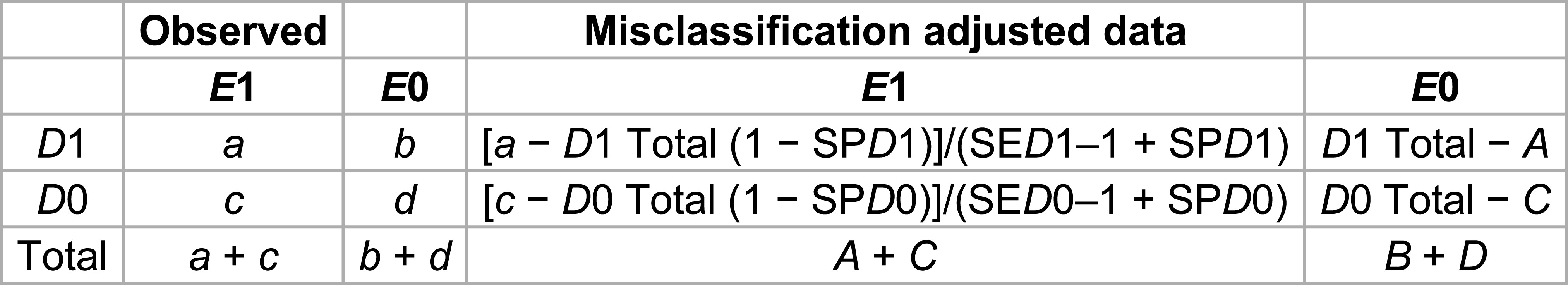

Here, SED1 and SED0 denote the sensitivity in cases and non-cases, respectively, while SPD1 and SPD0 represent specificity in cases and non-cases, respectively.

**eTable 1.**
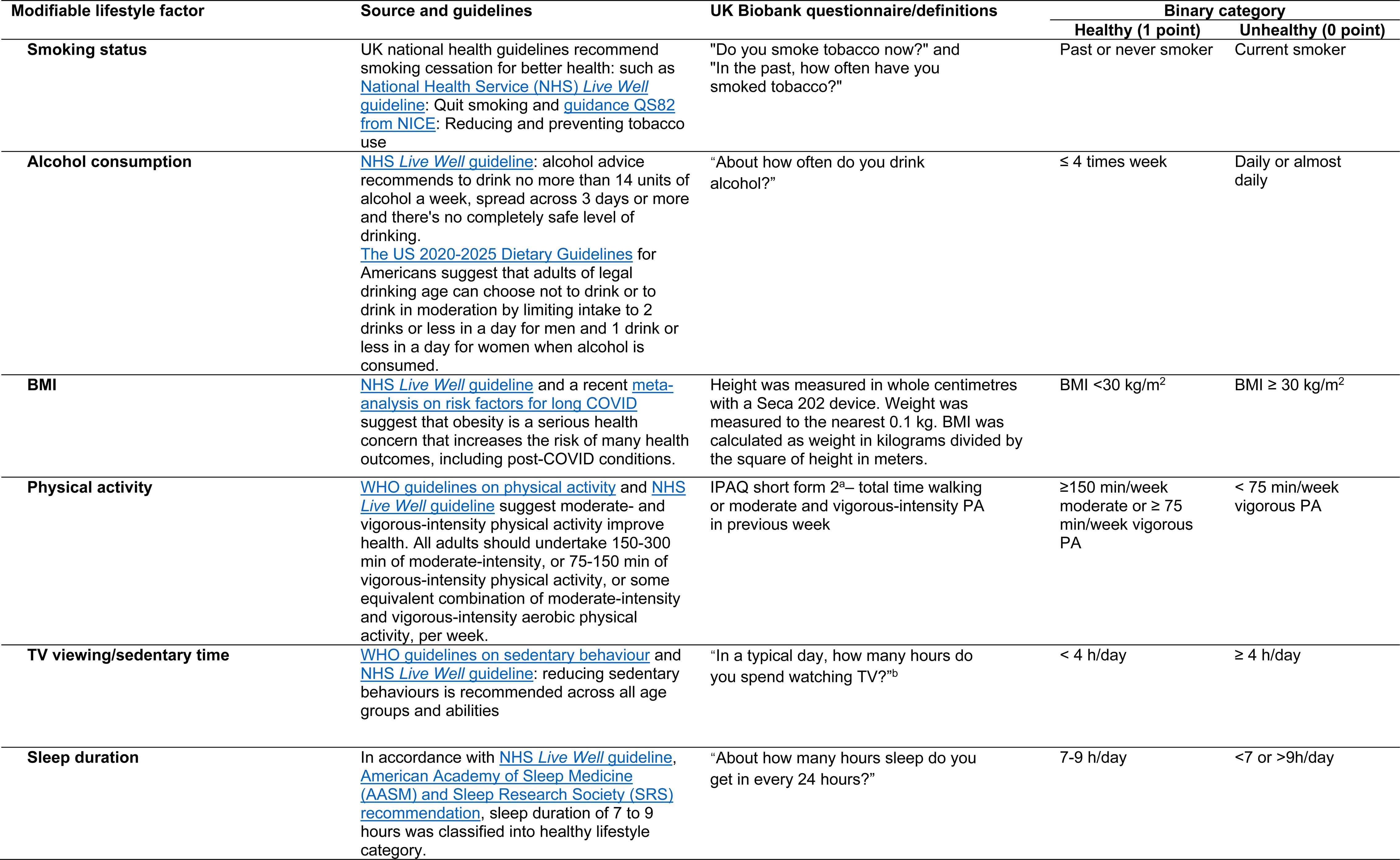
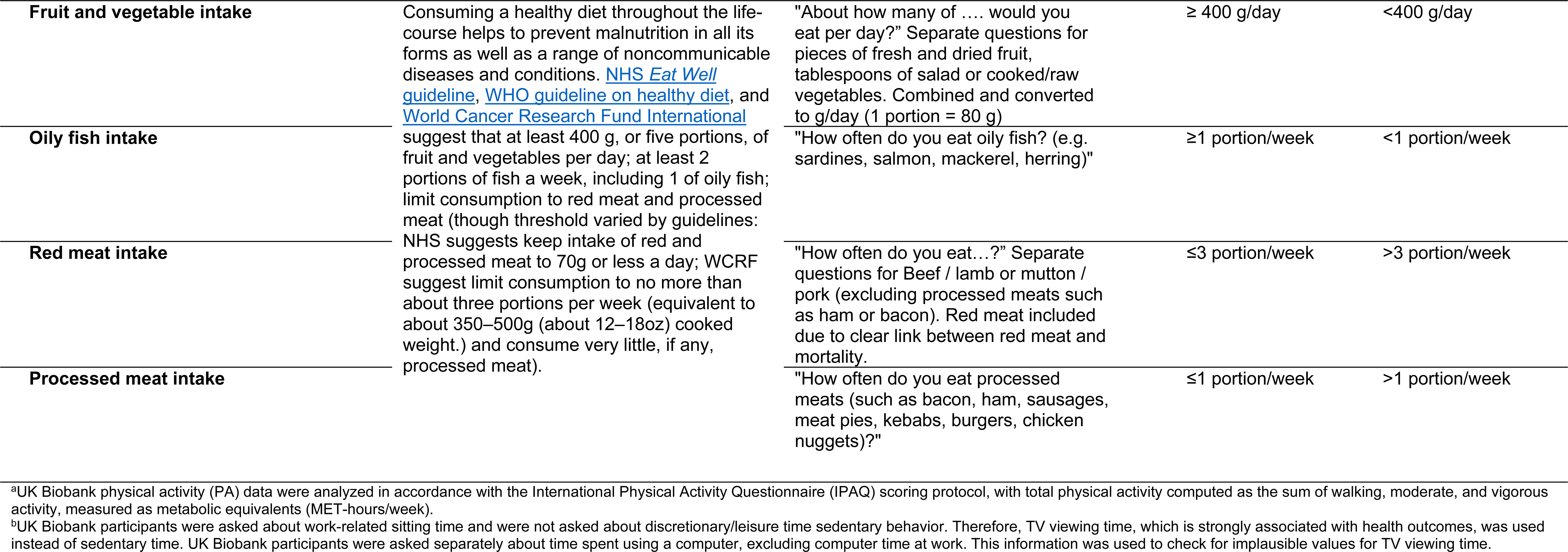
Detailed definitions on measurement and classification of lifestyle factors.

**eTable 2.**
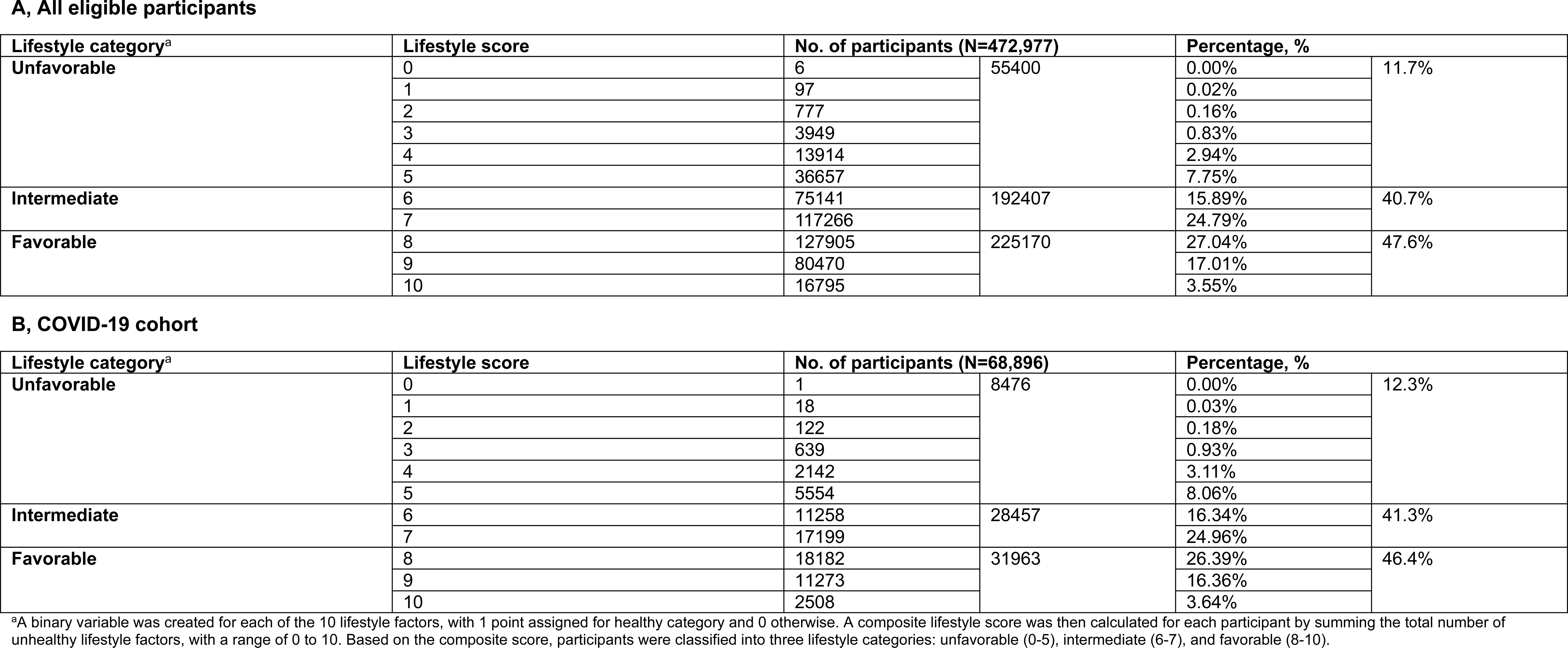
Distributions of lifestyle score and categories.

**eTable 3.**
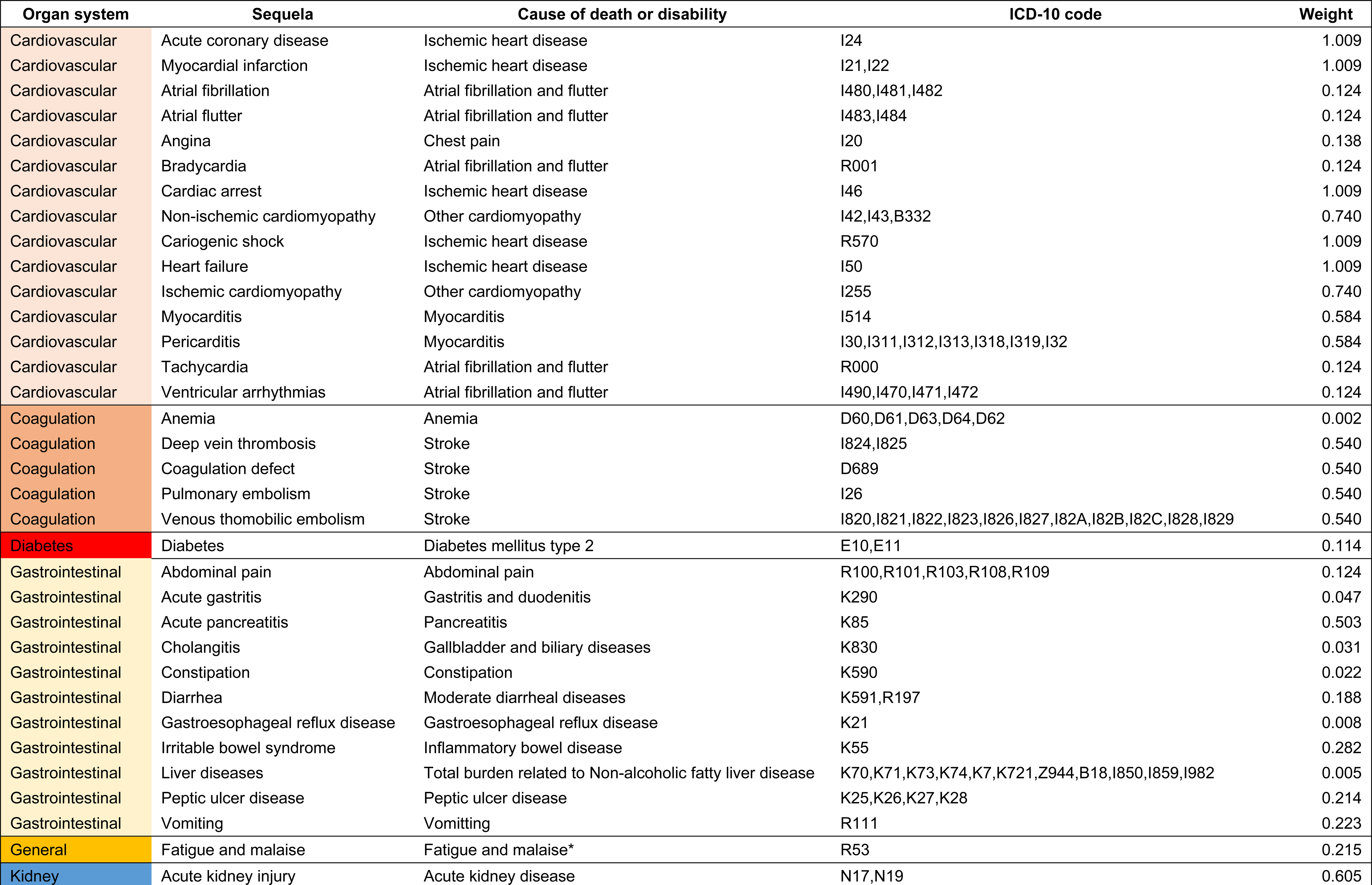
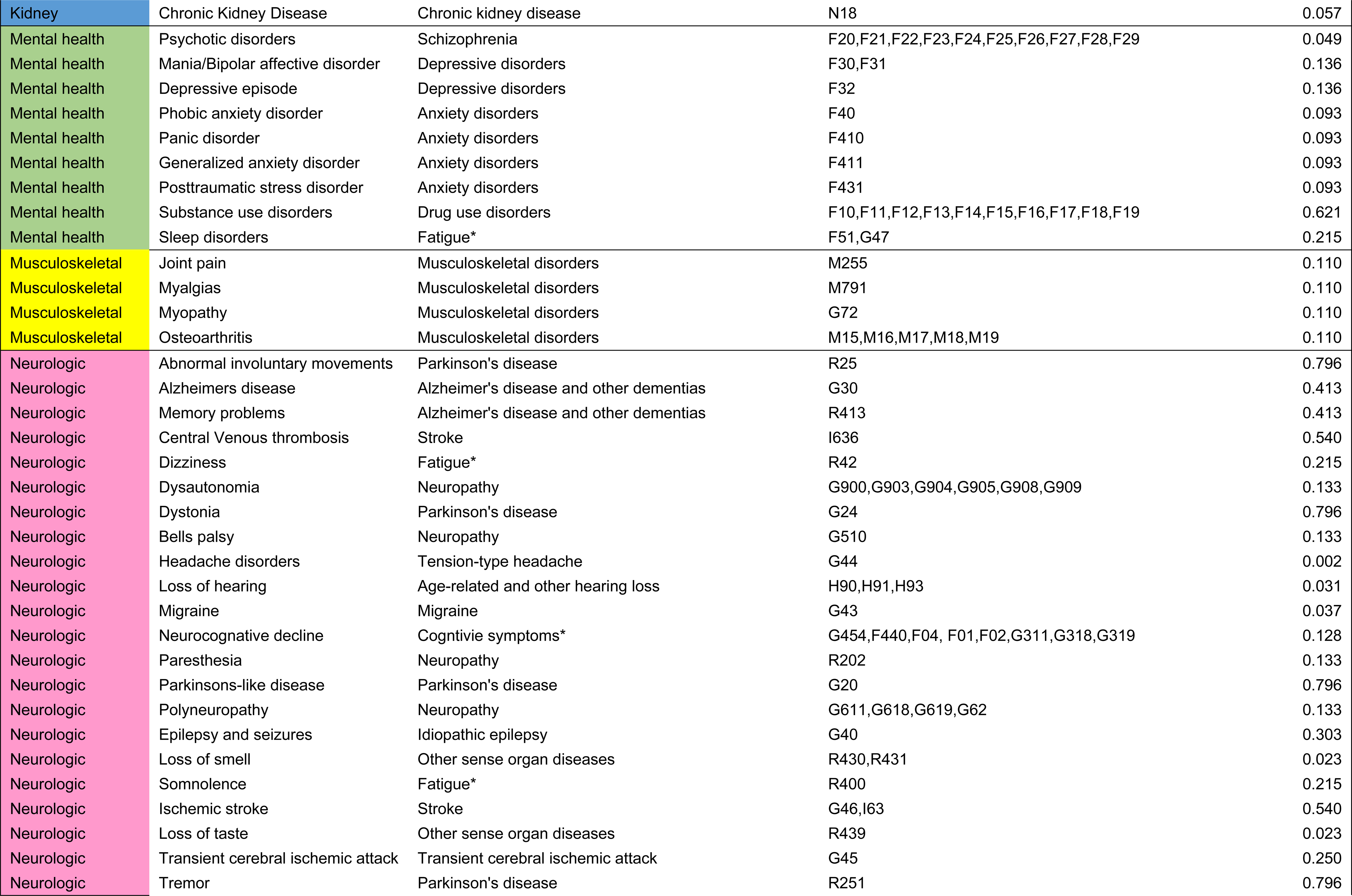
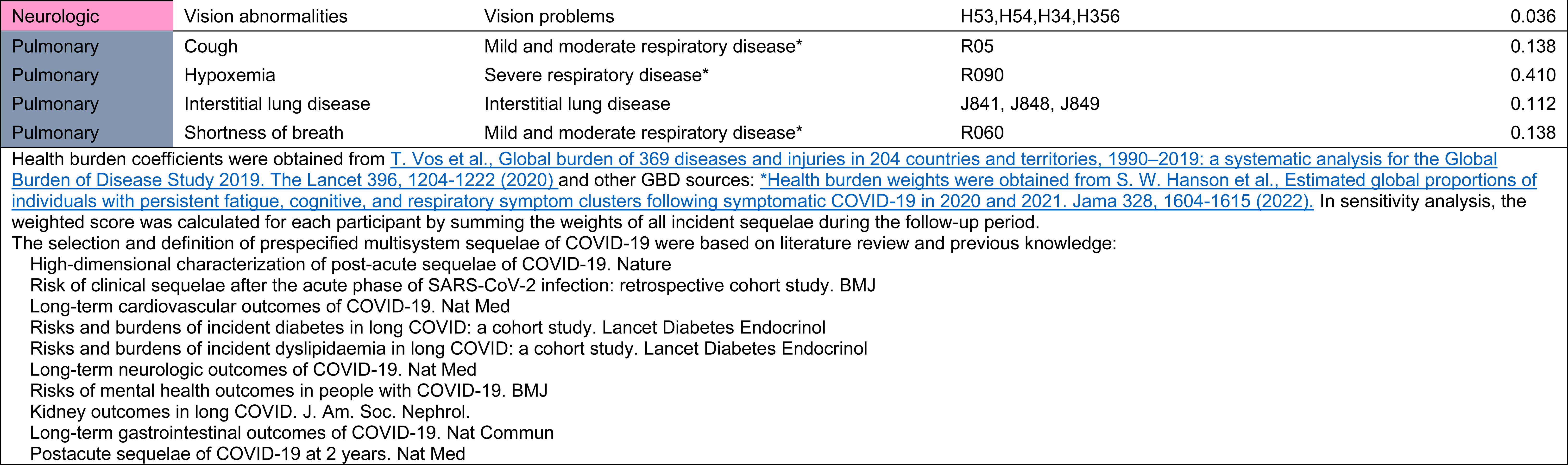
Definitions and weights of multisystem sequelae.

**eTable 4.**
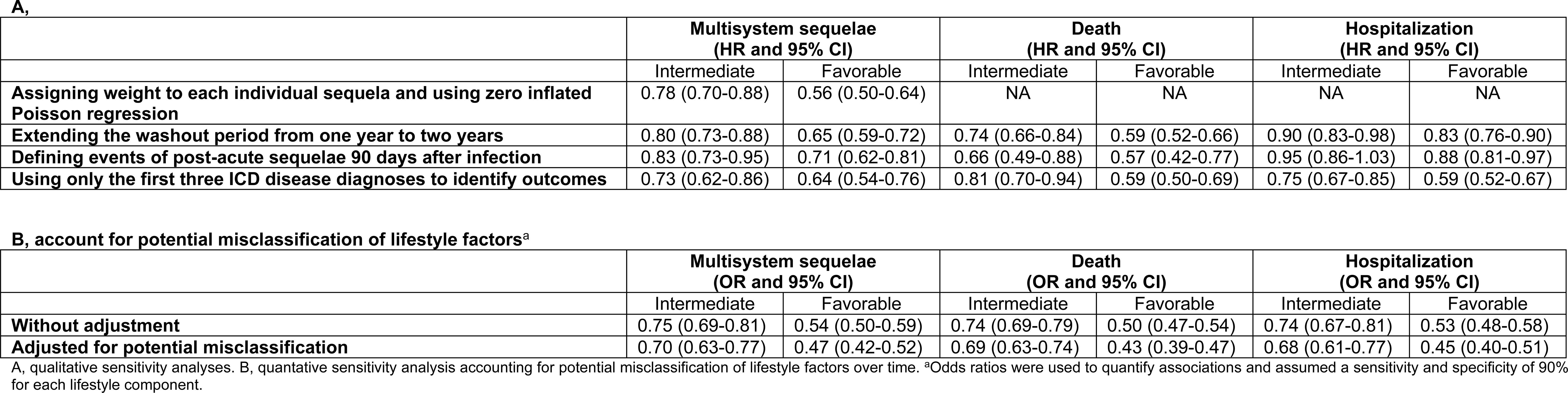
Sensitivity analyses of the risk of composite multisystem sequelae, death, and hospitalization.

**eFigure 1.**
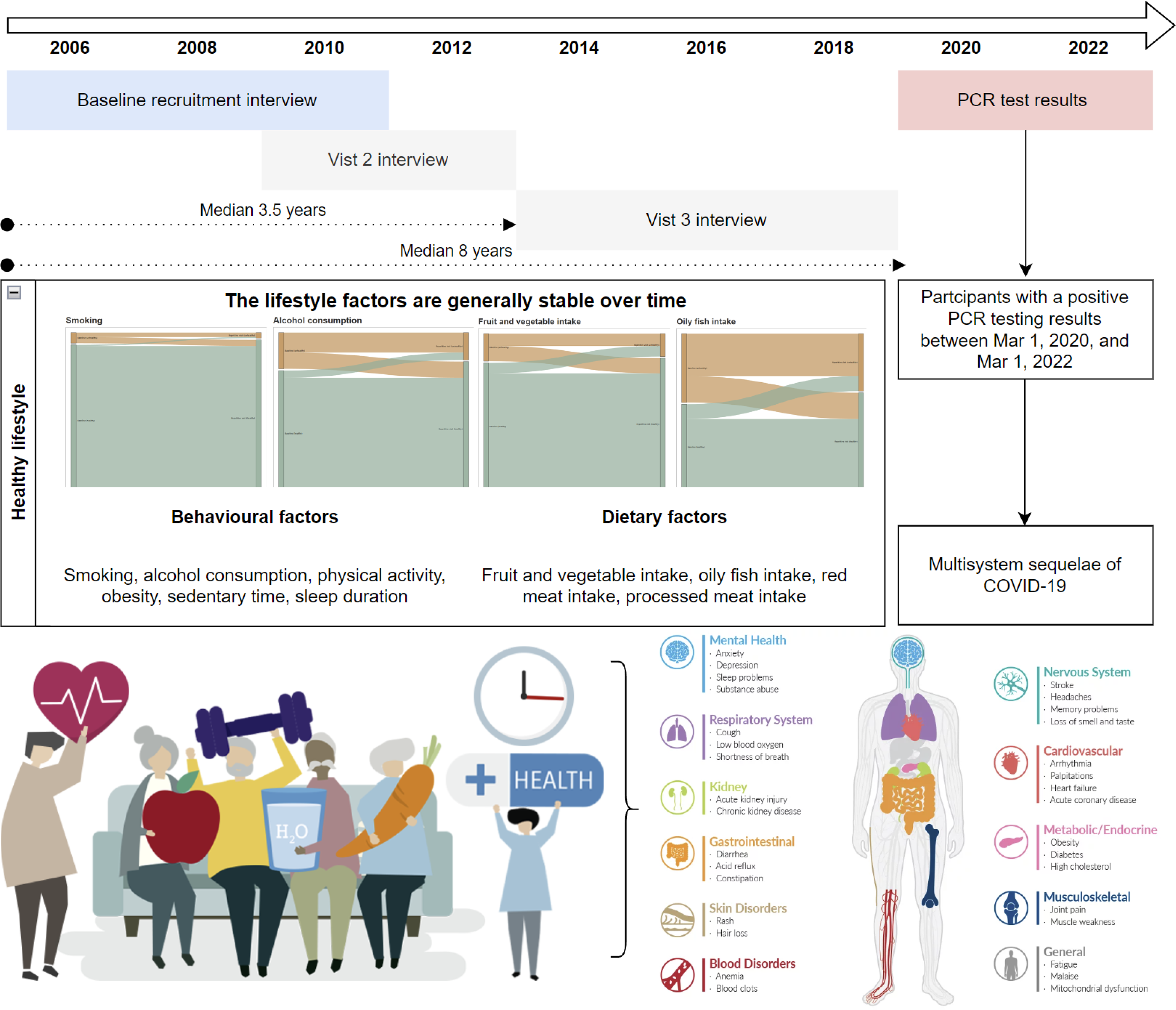
Study design, cohort construction, and timeline.

**eFigure 2.**
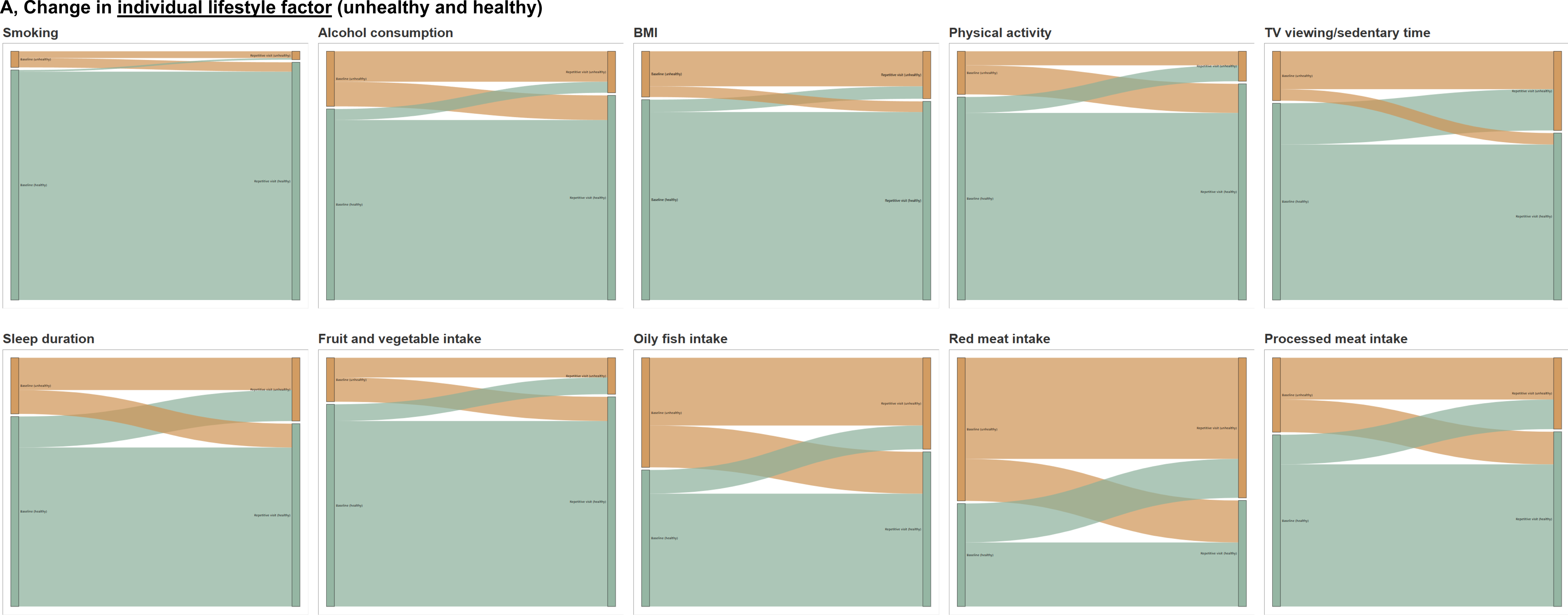
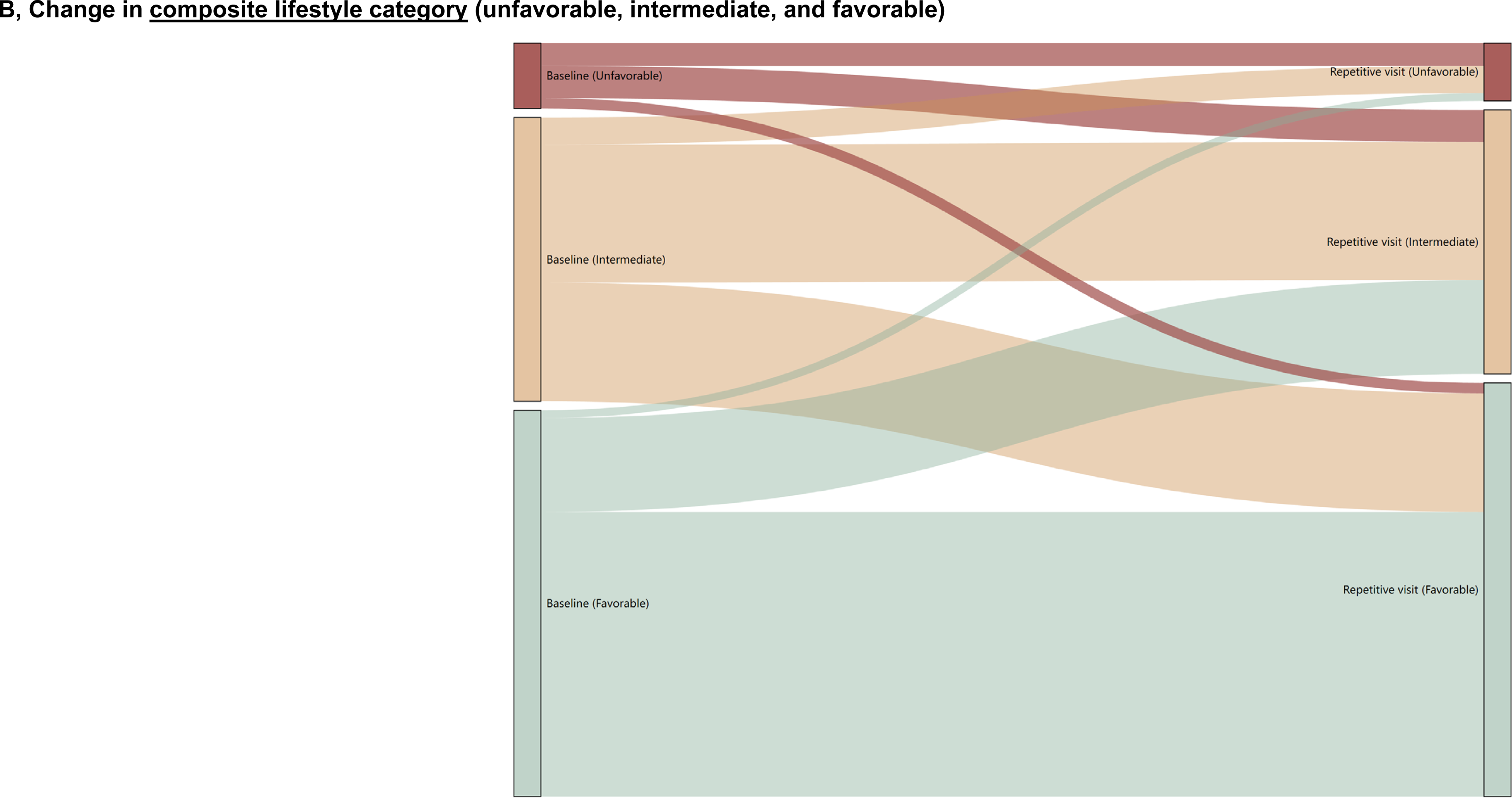
Change in lifestyle factors between baseline and the latest repeat assessment for those who undertook both visits. A, Change in 10 individual lifestyle factor between baseline and the latest repeat assessment. B, Change in composite lifestyle category between baseline and the latest repeat assessment. Left column of each plot indicates baseline and right column indicates repeat assessment. For each individual lifestyle factor as binary variable, unhealthy category is in orange, favorable category in green. For each the composite lifestyle as categorical variable, unfavorable category is in red, intermediate category in yellow, and favorable category in green. 34.9% of participants with an unfavorable lifestyle, 48.6% with an intermediate lifestyle, and 73.7% with a favorable lifestyle at baseline remained in the same corresponding lifestyle category at the latest repeat assessment ∼8 years later (overall proportion of stable categories, 60.6%). The sample size for each factor varied by missing conditions (N=∼60,000).

**eFigure 3.**
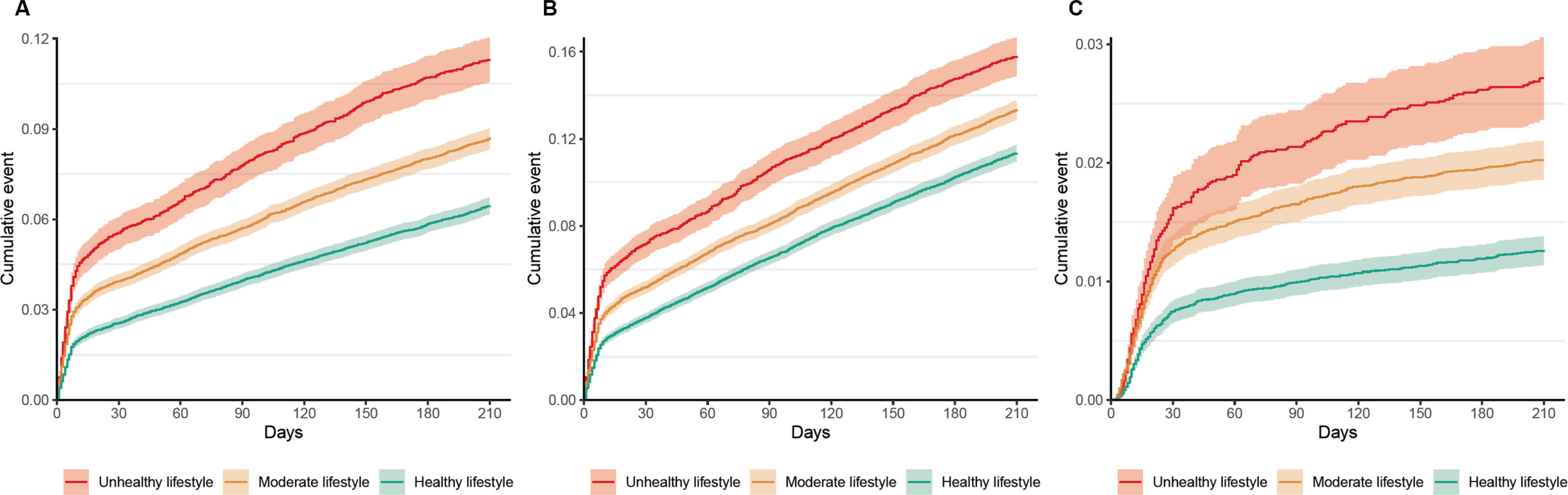
Cumulative incidence curves of composite multisystem sequelae, death, and hospitalization following SARS-CoV-2 infection. A, Composite multisystem sequelae. B, Death. C, Hospitalization. Outcomes were ascertained 0-210 days after SARS-CoV-2 infection. Event rates presented for the unhealthy lifestyle category (red), the intermediate lifestyle category (orange), and the favorable lifestyle category (green). The shadow of curves represents 95% confidence interval.

